# Development of an H&E on-block staining technique for collagen detection in cryo-fluorescence tomography imaging of frozen breast tissue samples

**DOI:** 10.1101/2024.12.20.24319432

**Authors:** Erin P. Snoddy, Tien T. Tang, Natalie Fowlkes, Thomas Huynh, Kari J. Brewer Savannah, Alejandro Contreras, Gregory Reece, Kristy K. Brock

## Abstract

**Background:** Hematoxylin and eosin (H&E) staining is widely considered to be the gold-standard diagnostic tool for histopathology evaluation. However, the fatty nature of some tissue types, such as breast tissue, presents challenges with cryo-sectioning, often resulting in artifacts that can make histopathologic interpretation and correlation with other imaging modalities virtually impossible. We present an optimized on-block H&E staining technique that improves contrast for identifying collagenous stroma during cryo-fluorescence tomography (CFT) sectioning.

**Approach:** In this prospective study, we embedded four breast specimens with confirmed ligaments from a bilateral mastopexy in an optimal cutting temperature block. Two of the samples were processed on a CFT imager and stained with our on-block staining protocol. In this protocol, hematoxylin was applied to the block face before being washed with deionized water. Eosin was then applied and washed with 95% ethanol. We then applied mounting medium and acquired images with a stereo-dissecting microscope and camera. Prior to staining, GFP fluorescence and white-light images were acquired with the CFT system to serve as a validation metric. The other two samples were sectioned on a standard cryostat and stained according to gold-standard H&E protocol. The resulting microscope slides were imaged with a digital slide scanner and viewed with Leica Imagescope software. An experienced pathologist evaluated both sets of images for qualitative comparisons.

**Results:** Pathologist evaluation confirmed that striations from on-block staining were qualitatively comparable with collagen tracks identified in gold-standard histology images. Furthermore, GFP images captured collagen autofluorescence, which aligned with the same structures identified by our on-block staining protocol.

**Conclusion:** Our on-block staining technique shows comparable visualization of collagenous structures at the mesoscopic level for fresh breast tissue samples. This technique improves tissue contrast and region of interest selection for histology during CFT imaging for analysis of the stromal architecture of the breast.

## Introduction

Hematoxylin and eosin (H&E) histology is widely considered to be the gold standard for diagnostic and surgical pathology in breast cancer. Correlative histopathology is important for validating findings obtained with various imaging modalities, such as magnetic resonance imaging (MRI) or computed tomography (CT). While these modalities utilize advanced imaging techniques that streamline numerous challenges in medical detection and diagnosis, it’s essential to have a reliable ground truth for assessing the efficacy and accuracy of their procedures. H&E histology plays a critical role in providing this grounding. Unfortunately, the high-fat content in breast tissue makes it a notoriously difficult tissue type to work with in histopathology labs. Adipose tissue does not freeze well at typical cryostat temperatures and is thus prone to smearing and tearing, often resulting in low-quality histology images.^1-3^ Various techniques help combat these difficulties, including freezing sprays, cooling the cryostat to lower temperatures, or acquiring thicker sections.^3^ However, the challenges associated with processing fat still make it difficult to study other heterogeneous components, including the collagenous structures comprising the breast tissue’s ligaments. We propose a new staining technique to identify collagen in breast tissue that involves the application of H&E stains directly to the embedded tissue block face for improved region of interest selection for histology. This technique is simple and fast and leaves the larger 3D volume of the specimen intact for further experimentation and analysis. This manuscript describes a comparison study between gold standard H&E staining and our novel on-block staining protocol for fresh breast tissue.

## Methods

### Breast Specimen Preparation

For this secondary analysis prospective study, we obtained four excised breast specimens from a female patient’s bilateral mastopexy procedure (two from the right breast and two from the left). This tissue was obtained from a consented participant enrolled in our institutional review board-approved study (PA16-0364). While this study took place in December 2023, optimization using 12 other breast specimens occurred between October 1^st^ and December 31^st^, 2023. All patients included in this study provided written consent. Exclusion criteria included ages younger than 18 years and older than 80 years, patients who underwent breast surgical procedures resulting in parenchymal scarring, and patients who were treated with partial or whole breast irradiation.

Following the excision of the breast specimens, approximately 2 inches long, an experienced breast reconstructive surgeon (G.R., 34 years of experience) identified and marked a prominent ligament in each specimen with a surgical suture knot. All specimens were flash-frozen in a dry ice/hexane bath to preserve their original shape and structure. Once frozen, each specimen was cut in half to ensure ligament presence in each tested sample. For each comparison experiment, two small samples (from left and right breast) were embedded in an Optimal Cutting Temperature (OCT) block for cryo-preservation using molds from a cryo-fluorescence tomography (CFT) imaging system (Xerra, EMIT Imaging, Baltimore, MD). Squid ink spaghetti noodle fiducial markers were placed in the mold to retain orientation. The OCT block was placed in a −20°C freezer until frozen before being mounted on the vertical sliding rail in the Xerra cryo-chamber for sectioning. The other two specimen halves were embedded in an OCT compound for conventional cryo-sectioning. An overview of the study workflow is shown in Figure 1.

**Figure 1:**
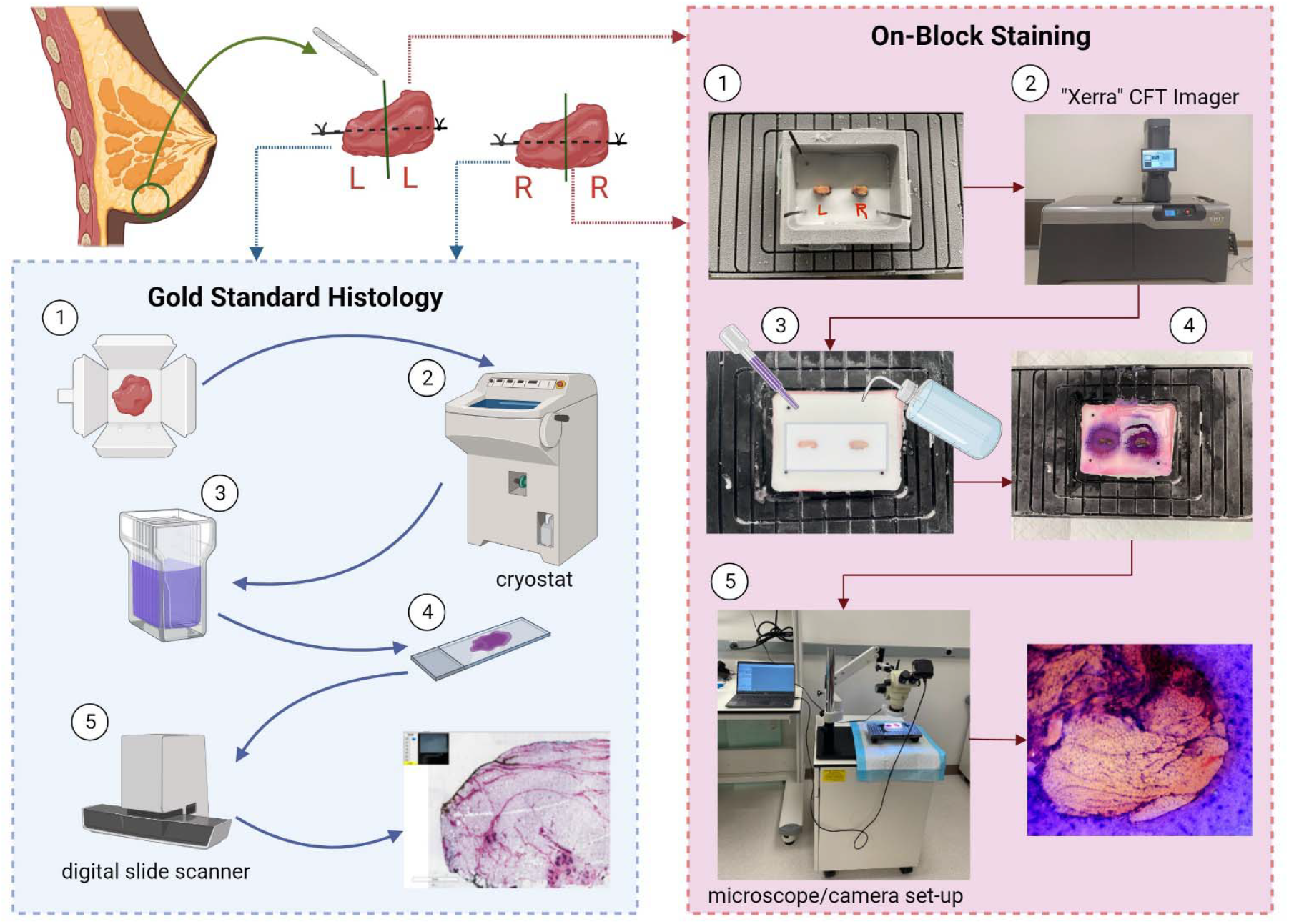
Illustrative workflow describing our experimental design. Created with BioRender.com

### CFT Imaging Protocol

All white-light and fluorescence images were acquired on the CFT imager using a 12-megapixel camera. Fluorescence imaging was done with a GFP filter (excitation: 470 nm, emission: 500 nm), and images were obtained at 50 µm intervals. A CFT imaging protocol was created to consecutively capture a white-light image, followed by a fluorescence image. The block was then automatically sectioned before the imaging protocol was repeated until the entire block was processed. Acquired images were reconstructed and registered using a multi-modality image post-processing suite (Invicro, VivoQuant, Needham, MA).

### H&E On-Block Staining Protocol

At planned sectioning intervals indicated by the suture knots, the block was removed from the CFT chamber for staining at various depths. A layer of medical gauze was placed over the surface of the block, and sufficient hematoxylin was applied with a disposable pipette to cover the entire tissue face. We allowed the block to sit for 60 seconds to enable the penetration of the stain into the tissue. The block was tilted at a slight angle, and a wash bottle with deionized water was used to wash the stain off the block, mimicking the rinsing step associated with standard protocols. A new piece of medical gauze was used to apply the eosin counterstain in the same manner as the hematoxylin. We let the stain sit for 30 seconds before tilting the block and washing it with 95% ethanol three times.

The timing was selected following experiment-based optimizations to enable the specific goals of this work. Increasing staining time showed a darker and more saturated stain, which could be adopted if preferred (Fig 2). A blueing step was attempted, as is the standard for H&E staining; however, we found no substantial differences with its incorporation, so it was not included in the final protocol.

**Figure 2:**
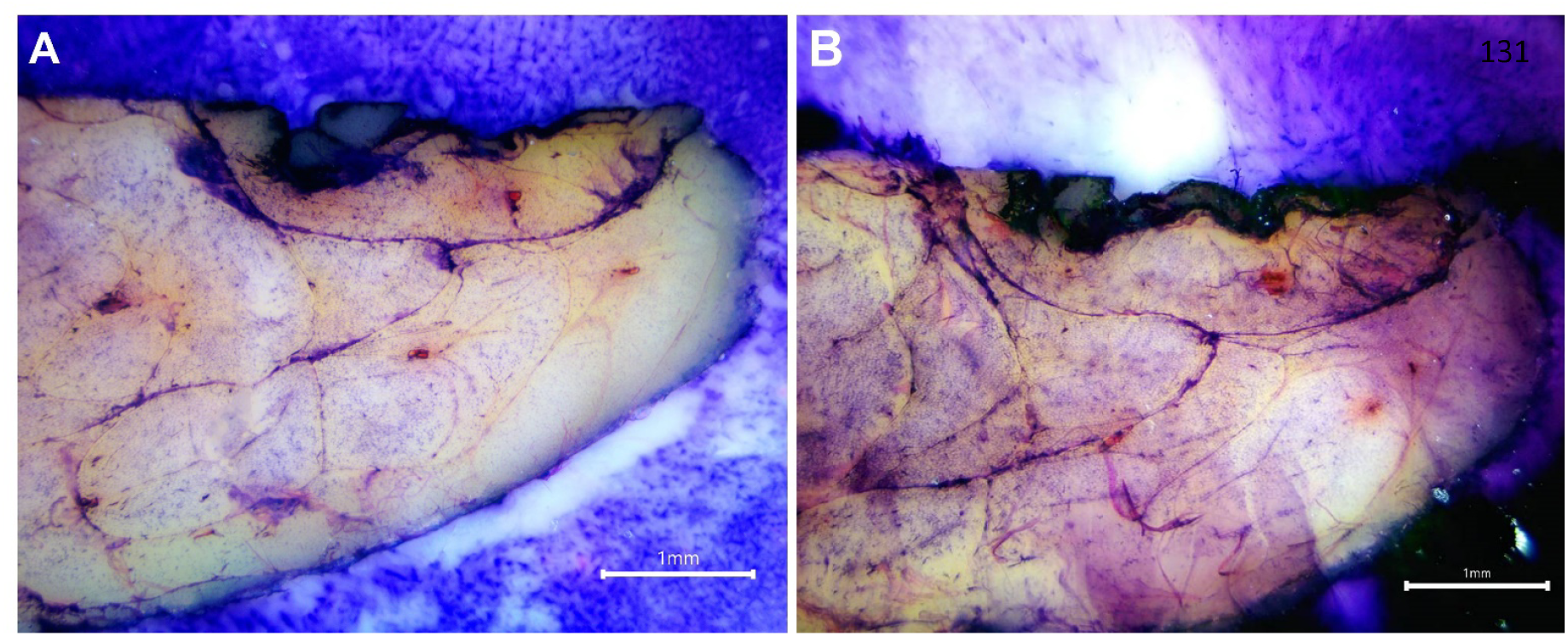
Visual representation of the effect of different staining times at 2x magnification. **(A)** Light staining; Protocol: 60 seconds hematoxylin, 30 seconds eosin. **(B)** Dark staining; 120 seconds hematoxylin, 60 seconds eosin.

### Microscope Imaging

Images were acquired using a stereo boom microscope (Omano, OM2300S-V1), a digital microscope camera (Summit SK2-14X), and a ring light. ToupView imaging software (ToupTek Photonics, Zhejiang, China) was used to acquire and process the images. We applied mounting medium (SouthernBiotech, Fluoromount-G®) to the tissue to improve image clarity. Areas of interest were photographed at 1x, 2x, 3x, 4x, and 4.5x using the auto-exposure function. These areas were informed by pictures of the gross specimen that showed the suture knots’ presence, indicating the location of a confirmed ligament. Other areas were informed by the fluorescence image taken by the CFT device prior to block removal from the cryo-chamber. After imaging, the OCT block was refrozen. After repeated experiments, we concluded that the block could stay under the microscope/ring light for approximately 30 minutes before the surface melted to a degree that impacted the experiment. We waited approximately 15 minutes for complete refreezing before resuming the CFT imaging study, allowing the CFT imager to continue slicing the block until another stain-free surface was exposed. We found the stain depth penetration to be roughly 0.82 mm +/-0.21 mm, or 16 slices (see Supplemental Materials for analysis).

### Gold Standard Histology Imaging

The other two specimen halves underwent gold-standard H&E staining for frozen sections. The halves were embedded directly onto the chuck (in the transverse direction to reflect the sample block orientation). 35 µm tissue sections were cut on a cryostat (CM3050 S, Leica Biosystems, Deer Park, IL) and transferred to charged microscope slides. These slides were then stained using a routine histology protocol. Slide images were captured with a digital slide scanner (Aperio AT2, Leica Biosystems, Deer Park, IL).

### Staining Technique Assessment

An experienced pathologist (N.F., 14 years of experience) evaluated a series of images from gold-standard H&E-stained frozen breast tissue at 1x, 2x, and 4x, and on-block images at 1x and 4x. The images were curated by excluding images with significant artifacts due to tissue tearing or folding.

## Results

Figure 3 depicts a qualitative comparison between a section of gold-standard H&E-stained slides containing breast tissue and an area of interest on the block that underwent our on-block staining technique. Qualitative evaluation by a board-certified pathologist (N.F.) determined that striations in the tissue highlighted by our staining approach are consistent with collagenous stroma and morphologically resembled collagen identified by conventional histology images. The images obtained at 1x magnification showed overall tissue architecture and morphology, while 4x magnification images provided enhanced resolution of structural detail, including delineation of adipocytes similar to that seen in traditional gold-standard histology images. All CFT white-light and fluorescence images were taken before the block was removed and stained, exhibiting the closest correlative match to the on-block-stained image (Fig 4). The GFP fluorescence images captured the autofluorescence of collagen within the breast ligaments (type I collagen is reported to have an emission band between 360 and 510 nm when excited at 337 nm).^4-6^ Areas of fluorescence align with structures highlighted in our on-block staining, suggesting that our technique accurately highlights collagen.

**Figure 3:**
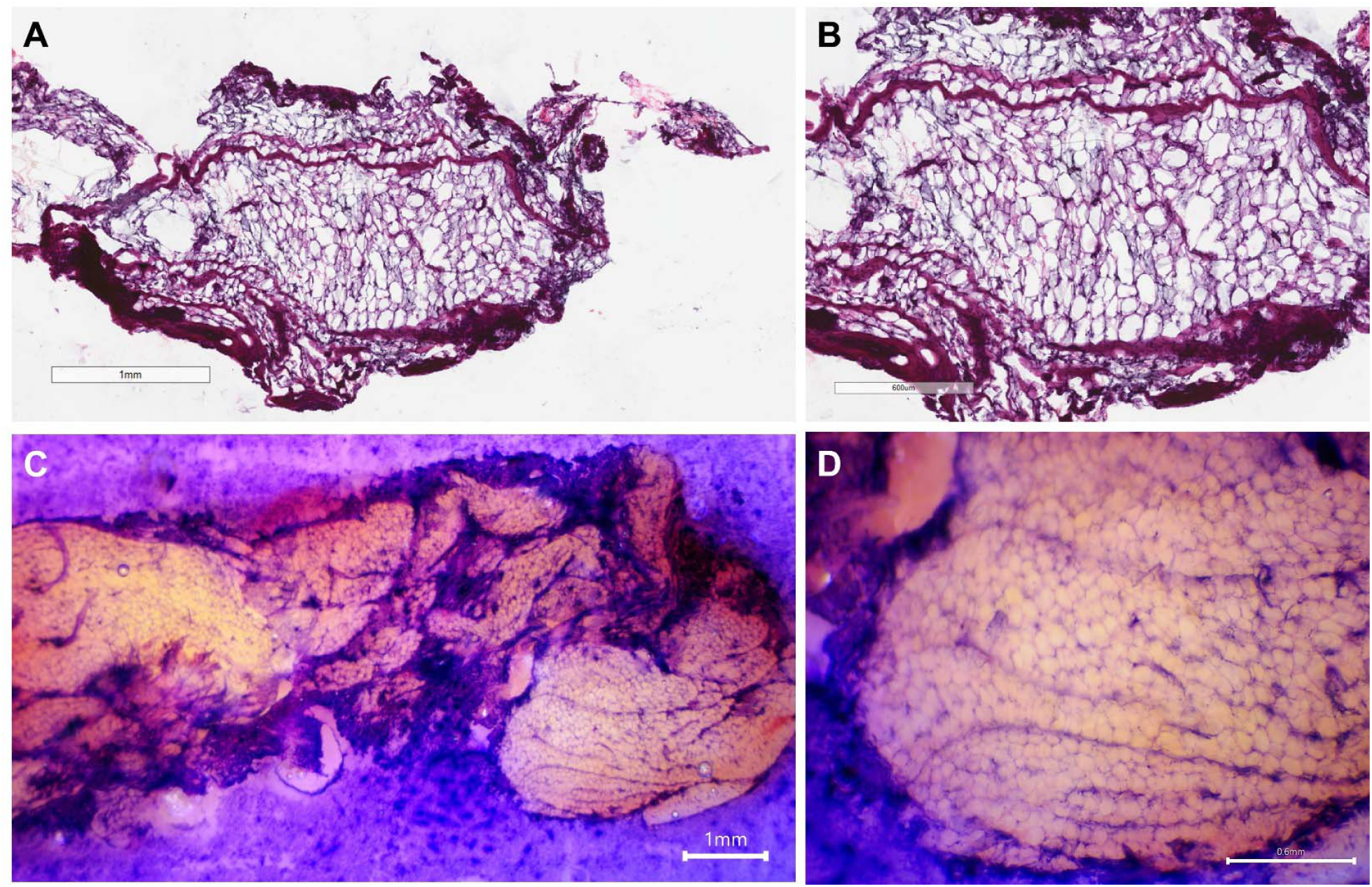
Qualitative comparison between gold-standard H&E (A-B) and on-block stained (C-D) breast tissue. **(C)** was obtained at 1x, **(A)** at 2x, and both **(B)** and **(D)** at 4x magnification. Adipocytes and collagen striation can be visualized with our on-block staining technique.

**Figure 4:**
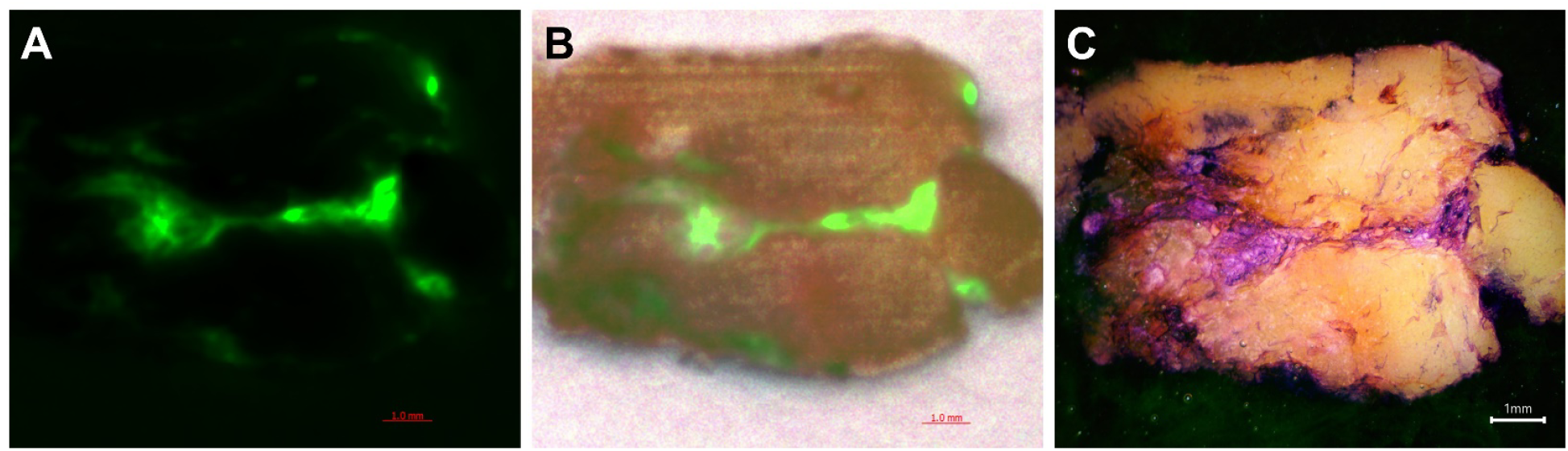
**(A)** CFT GFP fluorescent image. **(B)** CFT white-light-GFP fluorescent registration image. **(C)** Correlative on-block-stained image. All images were acquired at 1x magnification. The CFT images capture the autofluorescence properties of collagen. Areas of fluorescence in (A) and (B) co-localize to stained regions in the on-block image, further validating the staining technique.

## Discussion

In this study, we proposed a technique for obtaining H&E-stained images of breast tissue that highlights collagenous stroma for more sensitive detection of regions of interest in CFT imaging for research applications involving the breast. This technique provides improved structural collagenous detail at the mesoscopic scale and clear adipocyte delineation at the microscopic level. The morphology visualized in images obtained with our on-block staining technique resembles images obtained using a standard H&E protocol for frozen tissue.

The main advantage of this technique is that it provides an alternative methodology for the histological evaluation of breast tissue, a notoriously difficult tissue to section on a cryostat due to its fatty, non-aqueous nature. In addition, our protocol does not require tissue fixation, thereby preserving the structural nature with which the tissue was excised from the breast. It is also very fast; staining takes less than five minutes. A typical frozen H&E staining protocol takes at least 30 minutes, not including the time needed to section the tissue and complete slide transfer. We have shown that this imaging technique can be used in accompaniment with a commercially available CFT imager. Our protocol improves confidence in the detection of collagen, and the sample remains intact in the OCT block for further white-light and fluorescence imaging with the CFT device. This capability shows strong promise for future imaging-histology correlation studies. More specifically, the fluorescence in our CFT images shows a qualitative correlation with the areas highlighted by our on-block staining. In the future, we intend to use this quality as a validation metric to confirm that the collagen in our on-block-stained histology images are, in fact, breast fascial ligaments.

Much like how standard microscopy commonly calls for the use of mounting medium to improve optical clarity, we, too, have included mounting medium in our protocol to improve discernability under the microscope. Before this inclusion, our earliest microscope images featured a “droplet” artifact effect from the residual water/ethanol from the rinsing step (Fig 5A). We found that applying more ethanol so that the entire area of interest is immersed in liquid was sufficient to improve image quality and observation. However, ethanol dried on the tissue very quickly, leaving little time to operate the microscope for imaging. After repeated experiments, we determined that using an aqueous mounting medium, Flouromount-G, improved image quality without moving or drying out as quickly (Fig 5B).

**Figure 5:**
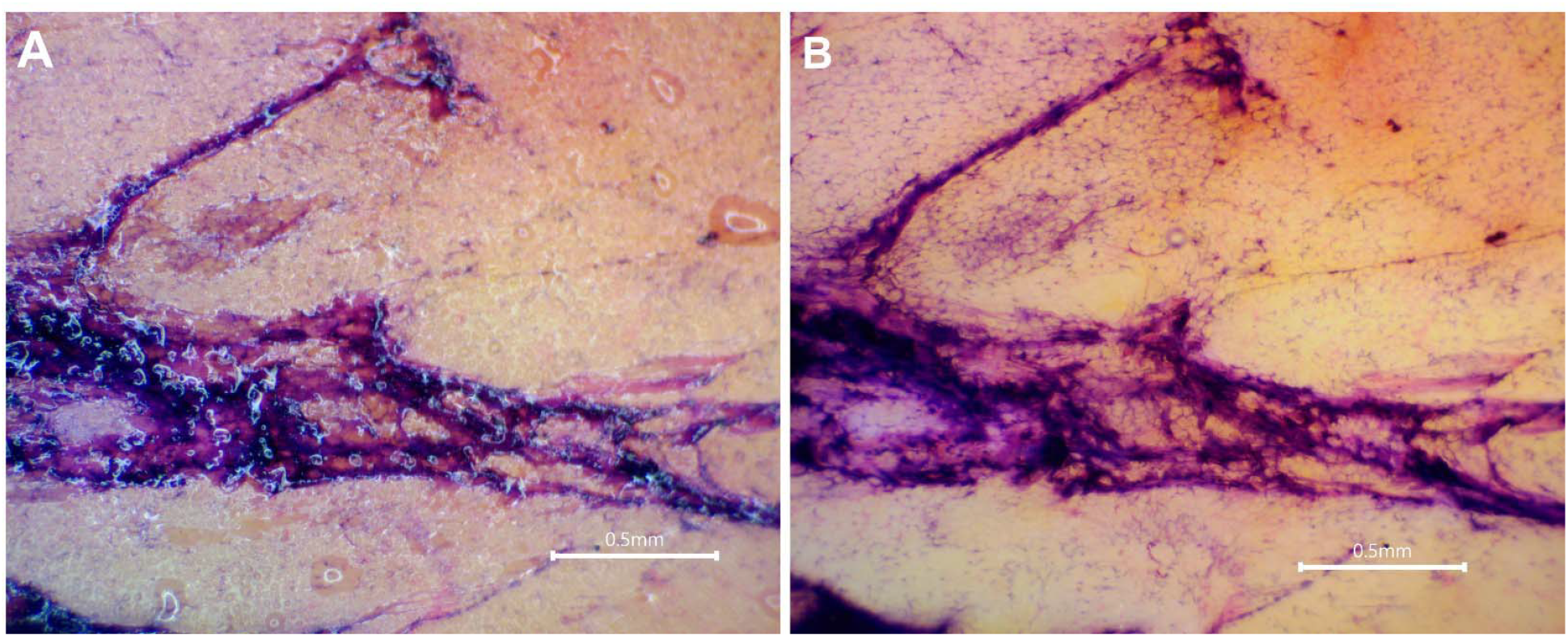
Visual representation of the effect of mounting medium. **(A)** Ethanol droplet artifact. **(B)** Revised protocol using mounting medium.

While useful for our group’s specific needs, we recognize that our technique does have limitations. Most notably, this includes the microscope’s limited imaging capabilities. Because of our OCT block’s large, opaque nature, we are confined to using a stereo-dissecting microscope to image our samples after on-block staining. The limited magnification and resolution make observing tissues at the single-cell and sub-cellular levels challenging. This could possibly limit our technique’s suitability for other research applications. Conducting on-block staining on a small, relatively flat tissue sample before sectioning the tissue to microns on a standard cryostat could feasibly work. However, it would likely require more experimentation. Furthermore, while we have only demonstrated this technique on human breast tissue, more optimizations would likely be needed for other tissue types or species, as well as for diseased tissue with significant necrosis, inflammation, or fibrosis. Finally, inherent variability introduced by hand-staining procedures such as our on-block staining protocol could result in inconsistencies limiting robust downstream quantitative or correlative analysis.

## Conclusion

We have optimized an H&E protocol that is compatible with a CFT device for staining large sections of breast tissue. Our protocol enhanced contrast and delineated collagen and adipocytes in frozen OCT-embedded breast tissue during CFT imaging, demonstrating detailed anatomy of the stromal architecture of the human breast. This technique shows great promise for facilitating multi-scale correlation from pathology to anatomical imaging.

## Data Availability

Any data is available upon request in compliance with institutional IRB requirements.

## Acknowledgments

The authors thank all the participants for their donation to this study. We also thank Mary Catherine Bordes and Sara Hull for their invaluable efforts in recruiting eligible women for this work and Charles Kingsley and Jennifer Meyer for their support with the Xerra CFT imager machinery. Finally, we thank Dr. Mingjie Wang, MD, for his assistance with breast ligament determination. This research was supported by the National Institutes of Health/NCI under award number P30CA016672 and the Small Animal Imaging Facility Core, the National Institute of Biomedical Imaging and Bioengineering of the National Institutes of Health under award number R01EB032533, and the Image Guided Cancer Therapy Research Program at The University of Texas MD Anderson Cancer Center through a generous gift from the Apache Corporation. The authors have no relevant conflicts of interest to disclose.

