## Supplemental Figure 1 for "Development of an H&E on-block staining technique for collagen detection in cryo-fluorescence tomography imaging of frozen breast tissue samples"

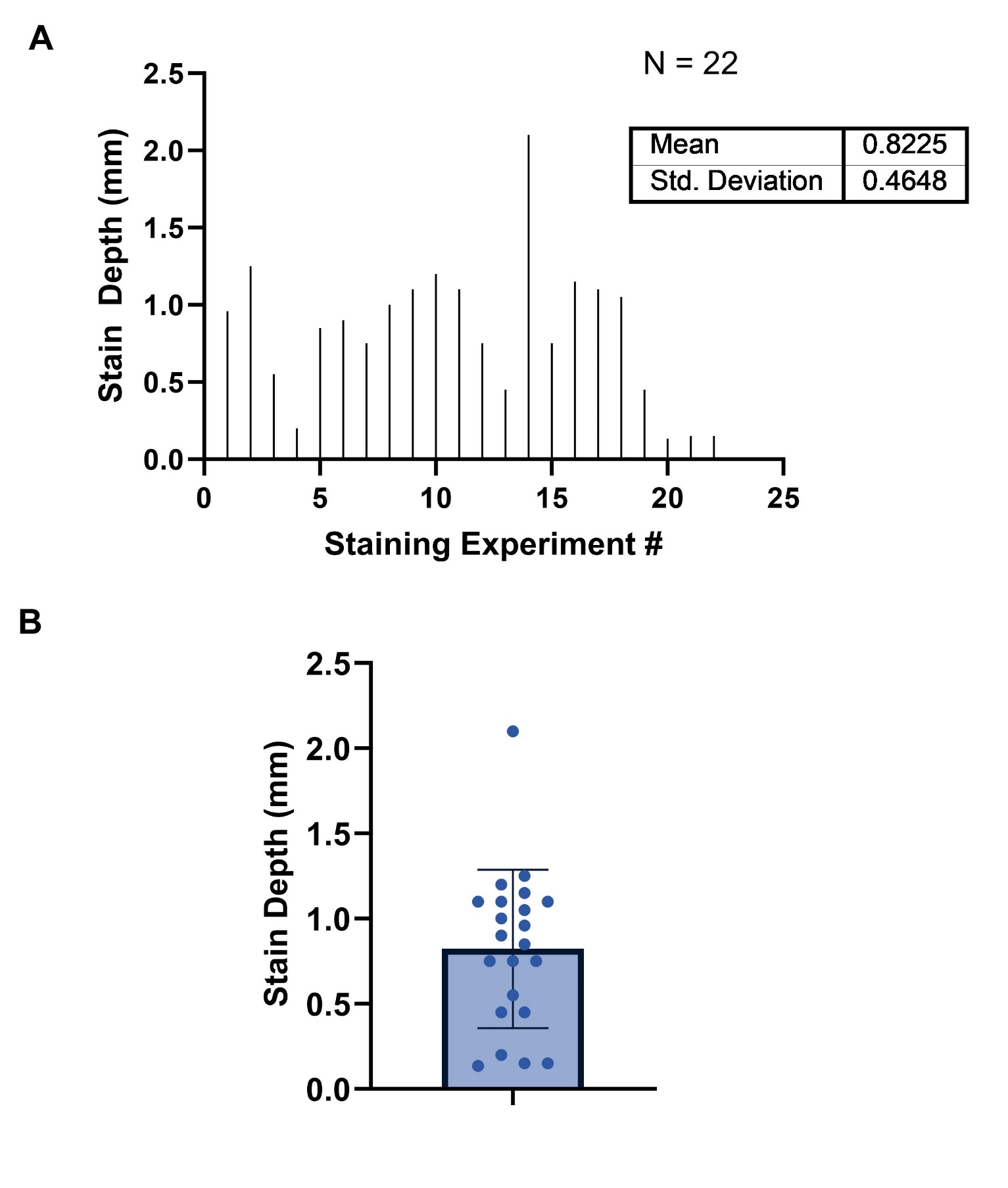

**Figure S1:** The reported stain penetration depths for 22 optimization experiments. After staining the block-face, the sample was returned to the cryo-chamber, and sectioning continued. Each successive 50-µm slice was counted until all the stain was removed and a new block-face was revealed. Some samples underwent multiple staining trials. A one-sample t-test was performed to compute descriptive statistics. **(A)** is a visual representation of the data in the form of a XY plot. Created with GraphPad Prism.
